# Clinician Discourse on Ambient AI Scribes: A Reddit-based Topic Modelling and Sentiment Analysis

**DOI:** 10.64898/2026.04.26.26351798

**Authors:** Ravi Shankar, Qian Xu

## Abstract

**Background:** Ambient AI scribes are rapidly entering clinical workflows, yet end-user perspectives remain underrepresented in the peer-reviewed literature. Online clinician communities offer an unfiltered window into adoption barriers, perceived benefits, and product-level concerns.

**Objective:** To characterise themes and sentiment in clinician discourse on ambient AI scribes across professional Reddit communities.

**Methods:** We scraped posts from ten clinically oriented subreddits using twelve AI scribe related queries via the public Reddit JSON API. A two-tier keyword filter retained posts mentioning at least one AI scribe term and one clinical or workflow term. Texts were embedded with all-MiniLM-L6-v2, reduced via UMAP, clustered with HDBSCAN, and labelled using BERTopic with c-TF-IDF keyword extraction. Noise topics matching predefined off-topic patterns (for example, residency match, finance) were removed. Themes were assigned concise labels via Claude Sonnet 4. Sentiment was classified per post using cardiffnlp/twitter-roberta-base-sentiment-latest.

**Results:** After filtering, 176 unique relevant posts from seven active subreddits were retained, with r/FamilyMedicine (n = 64) and r/healthIT (n = 34) dominating. BERTopic produced 12 coherent themes spanning workflow integration, vendor comparison (DAX, Heidi, Freed, Abridge), HIPAA and privacy, mobile and device use, templates and formatting, and research versus clinical use. Overall sentiment was 61.4% neutral, 21.6% positive, and 17.0% negative. The most net-positive theme was DAX/Nuance/AI tools (about 55% positive); the most net-negative were charting fatigue and the freed-AI-scribes discussion thread (about 37 to 40% negative). Engagement (median upvotes and comments) was highest for tool-comparison and pricing themes, indicating salience of practical adoption questions.

**Conclusions:** Clinician sentiment toward ambient AI scribes is cautiously favourable but dominated by neutral, problem-solving discourse. Vendor selection, cost, HIPAA compliance, and EHR integration are the most actively debated issues. These insights can inform implementation strategy, vendor benchmarking, and policy guidance for ambient documentation tools.

## 1. Introduction

Documentation burden is a leading driver of clinician burnout, with physicians spending one to two hours on the electronic health record (EHR) for every hour of direct patient care [1,2]. The cumulative effect on after-hours work, often referred to as pyjama time, has been quantified across specialties and is consistently associated with higher burnout scores and intent to leave clinical practice [3,4]. Ambient artificial intelligence (AI) scribes, which passively listen to the clinical encounter, transcribe it, and generate a structured note, have emerged as one of the most rapidly diffusing categories of clinical AI [5,6]. Commercial offerings such as Nuance DAX Copilot, Abridge, Suki AI, Heidi Health, and Freed have moved from pilot to enterprise deployment in less than three years, and several health systems now report measurable reductions in after-hours charting and improvements in clinician satisfaction [7,8,9].

Despite this momentum, the peer-reviewed evidence base remains thin and concentrated on a small number of vendor-supported pilots [10,11]. Real-world adoption questions, such as which tools work in which workflows, how they handle HIPAA and consent, what they cost in subscription terms, and how their notes integrate with Epic, Cerner, athenahealth, or eClinicalWorks (eCW), are debated daily on professional online forums but are rarely captured in formal studies [12,13]. Reddit communities for physicians, nurses, physician assistants, and health IT staff offer a uniquely candid corpus: discussions are voluntary, anonymous, and frequently include side-by-side comparisons of competing tools [14,15]. Prior work has used Reddit and other social media platforms to study clinician burnout, drug repurposing, vaccine hesitancy, and patient experience [16,17,18], demonstrating both the value and the methodological challenges of mining this signal.

Topic modelling has evolved rapidly in recent years. Early applications relied on Latent Dirichlet Allocation [19], which is sensitive to short texts and bag-of-words assumptions. Transformer-based approaches such as BERTopic combine sentence embeddings with density-based clustering and class-based TF-IDF, yielding more coherent and semantically meaningful clusters on small to medium corpora [20,21]. For sentiment, transformer models trained on social media text outperform classical lexicon-based approaches on noisy user-generated content [22,23].

We applied a transformer-based topic modelling and sentiment pipeline to clinician posts about ambient AI scribes on Reddit. Our objective was twofold: (i) to identify the latent themes structuring this discourse, and (ii) to quantify sentiment within and across themes. We hypothesised that themes would cluster around four practical concerns, namely workflow and time savings, tool comparison, privacy and compliance, and integration and accuracy, and that sentiment would be predominantly neutral, reflecting the problem-solving register of professional forums [16,24].

## 2. Methods

### 2.1 Data source and search strategy

We retrieved publicly available Reddit posts from ten clinically oriented subreddits: r/medicine, r/FamilyMedicine, r/healthIT, r/Psychiatry, r/physicianassistant, r/nursing, r/medicalschool, r/digitalhealthcare, r/EMR, and r/AIAssistants. Posts were collected via the public Reddit JSON search API (no authentication required; user agent declared as academic, non-commercial), an approach used in prior digital-health social-media studies [16,18]. Twelve search queries were used: AI scribe, ambient AI clinical, medical scribe AI, DAX Copilot scribe, Abridge, Nuance DAX scribe, Heidi Health scribe, Suki AI, AI clinical documentation, clinical notes AI, ambient documentation clinical, and AI charting. For each subreddit-by-query combination, up to 25 results were retrieved, with a 1.5 second delay between requests and a fallback retry on HTTP 429 responses. Comments were not collected for the present analysis.

### 2.2 Two-tier relevance filtering

Search APIs return many tangentially relevant posts [25]. To increase precision without manual screening, each post was required to satisfy a two-tier keyword filter applied to the concatenated title and body, an approach analogous to the dual-anchor strategy used in PRISMA-S literature searches [26]. Tier 1 (AI scribe specific) required at least one of: ai scribe, ambient scribe, ambient ai, medical scribe, ai documentation, ambient documentation, ai charting, ai note(s), ai clinical, dax, abridge, heidi, suki, copilot scribe, nuance scribe, dragon medical, voice ai, ai transcri-, scribe ai, scribe tool, ambient tool, or ambient listen. Tier 2 (clinical or workflow context) required at least one of forty-three terms covering EHR, clinician role, charting, accuracy, consent, privacy, HIPAA, cost, burnout, error, hallucination, and related operational concepts. Posts shorter than fifty characters after cleaning were also discarded. Duplicate post identifiers were removed.

### 2.3 Topic modelling

Documents were embedded using the all-MiniLM-L6-v2 sentence-transformer (384-dimensional output) [27]. Embeddings were reduced with UMAP (n_neighbours = 5, n_components = 5, min_dist = 0, metric = cosine, random_state = 42) [28] and clustered with HDBSCAN (min_cluster_size = 5, min_samples = 3, cluster_selection_method = expectation of mass) [29]. Topic representations were generated through BERTopic [20] using a CountVectorizer with English stop words and bigram support (min_df = 2), and a class-based TF-IDF (c-TF-IDF) transformer with frequent word reduction. Topic count was set to auto. To prevent residual off-topic clusters from contaminating the analysis, nine noise patterns reflecting recurrent off-topic threads (residency match, mental-health screening scales, loans and salary, baby/DNP transitions, profanity-laden rants, twitter cross-posts, generic job/PTO posts, school/loans/career posts, and PA programme posts) were applied; topics matching three or more keywords from any pattern were collapsed into the outlier class.

### 2.4 Theme labelling

For each retained topic, the top five c-TF-IDF keywords plus five exemplar posts were submitted to Anthropic’s Claude Sonnet 4 model with a structured prompt asking for a five to eight word clinical thematic label focused strictly on the AI scribe and documentation angle. LLM-assisted labelling has been shown to produce more interpretable topic descriptors than raw keyword lists in qualitative synthesis pipelines [30,31]. The returned label was used in all downstream visualisations alongside the raw keyword string for transparency.

### 2.5 Sentiment analysis

Each post was scored using the cardiffnlp/twitter-roberta-base-sentiment-latest model [22,23], fine-tuned on Twitter for three-class polarity (positive, neutral, negative). Texts were truncated to 512 tokens. The highest-probability class was assigned as the sentiment label and its softmax probability retained as the confidence score. A polarity score (+1, 0, −1) was derived for theme-level distribution analysis.

### 2.6 Visualisation and analysis

All figures were generated in Python (matplotlib, seaborn, wordcloud) [32]. Theme positioning was visualised via an intertopic distance map (UMAP-2D embedding of c-TF-IDF topic vectors) [20], bubble charts of positive versus negative sentiment, polarity violins, and engagement bar charts. Engagement metrics, namely median upvote score and median comment count, were computed per theme to identify the most discussed issues, after winsorising scores to [-10, 500] and comments to [0, 200] to limit outlier influence [33].

### 2.7 Ethics

Only public, non-personally-identifiable posts were analysed, in accordance with Reddit’s terms of service and prevailing guidance for online community research [14,34]. No usernames, geolocation, or off-platform identifiers are reported. The study was non-interventional and did not require institutional ethics review.

## 3. Results

### 3.1 Corpus characteristics

After two-tier filtering and de-duplication, 176 unique posts were retained from seven active subreddits (three of the original ten yielded no relevant content after filtering). The dominant communities were r/FamilyMedicine (n = 64; 36.4%) and r/healthIT (n = 34; 19.3%), followed by r/physicianassistant (n = 25), r/nursing and r/Psychiatry (n = 16 each), r/medicine (n = 15), and r/medicalschool (n = 6) (Figure 1). The skew toward family medicine reflects both the size of the subreddit and the disproportionate documentation burden in primary care, where ambient scribes have been most aggressively marketed [3,8].

**Figure 1.**
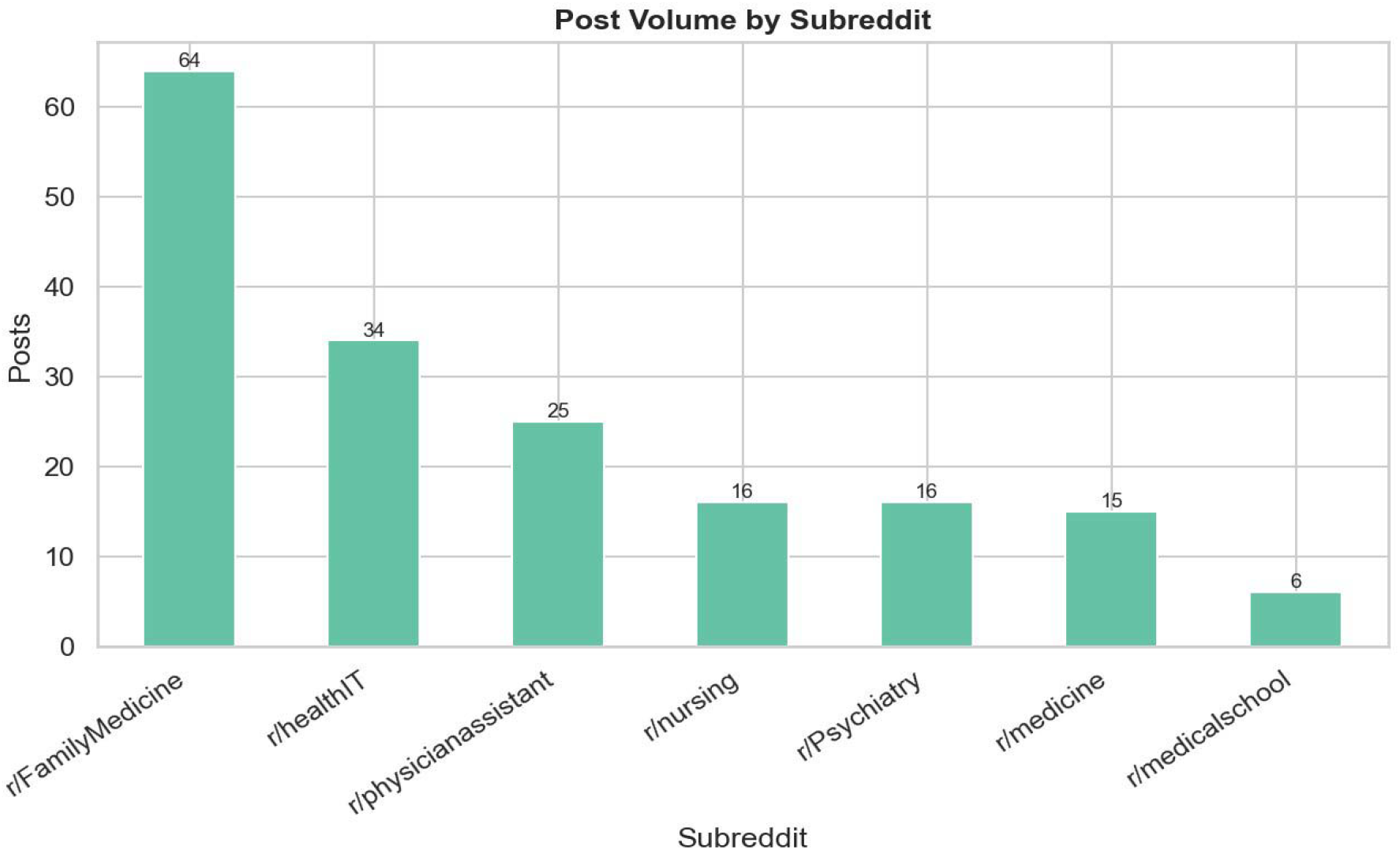
Post volume by subreddit after two-tier relevance filtering (n = 176).

### 3.2 Lexical landscape

The top thirty keywords across the corpus (Figure 2) were dominated by time, work, patients, day, free, care, health, clinic, scribes, year, job, charting, and help, a vocabulary that combines workflow and burnout language with explicit product references [3,4]. The prominence of free, month, and 99 reflects active discussion of subscription pricing (notably the Freed tier), while charting, templates, and visit point to the operational core of the documentation task. Epic appears at moderate frequency, indicating that EHR integration is a recurrent but not dominant concern [12].

**Figure 2.**
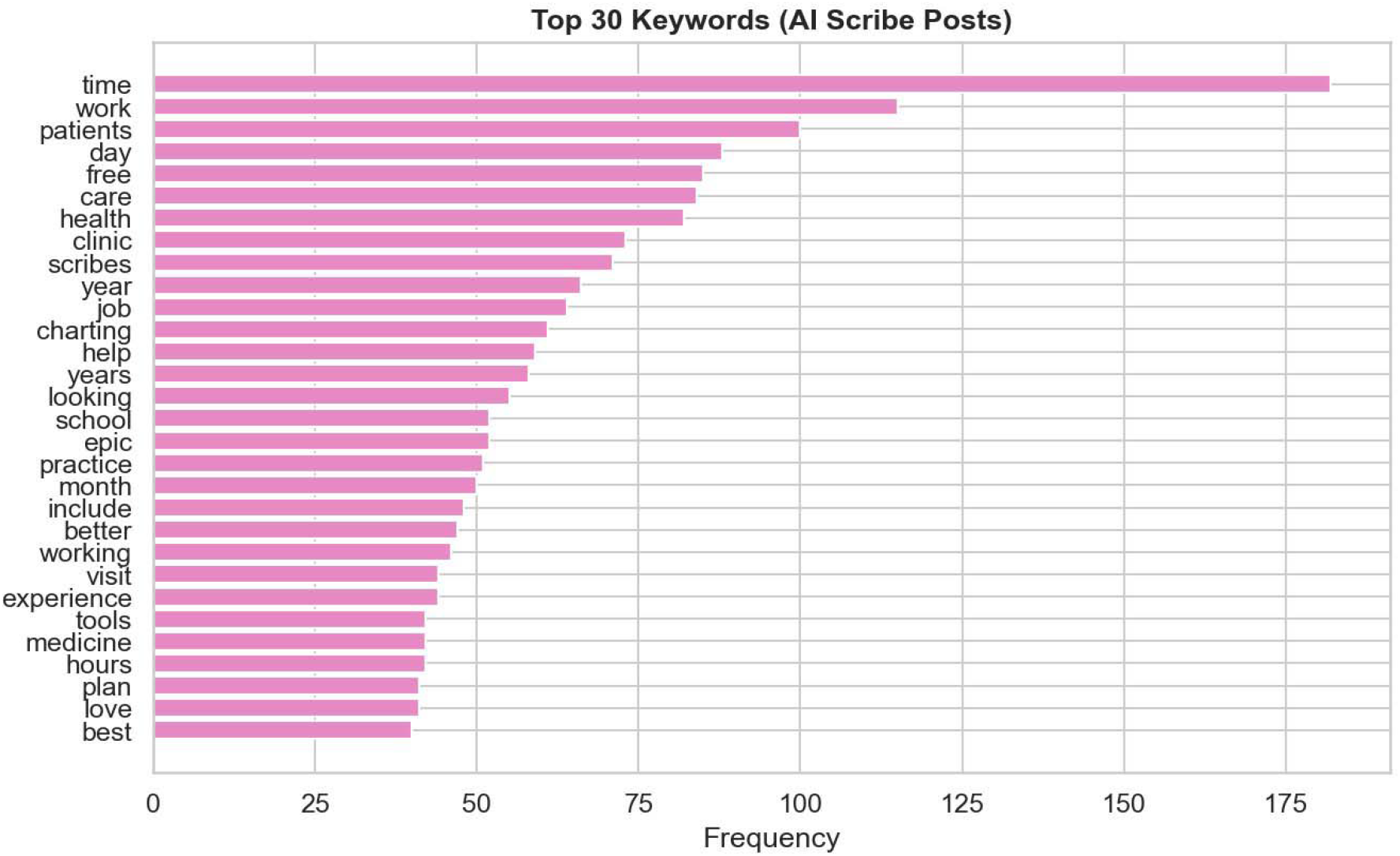
Top thirty keywords across all AI scribe posts. Time, workflow, and pricing terms dominate.

### 3.3 Latent themes

BERTopic yielded twelve coherent themes after noise removal, ranging from 5 to 16 posts each (Figure 3). The largest cluster (n = 16) centred on a recurrent thread about a flagged Freed AI scribe post, illustrating community moderation dynamics around vendor-related content [14]. Substantively, the themes can be grouped into five families: (i) vendor and tool comparison (DAX/Nuance/AI tools, scribe-tool/better-AI/integrated, twofold/templates/Heidi/formatting, DAX/sections/AWV/statements); (ii) cost and access (99/month/Freed/simple); (iii) compliance and trust (HIPAA/open-source/compliant); (iv) workflow and device context (phone/device/Chrome/hands, charting/end-of-shift/spending); and (v) clinical content and use case framing (specific/CI/language/changes/conditions, research/internet/ChatGPT, eCW/organisations/technology/products/workflow). This grouping is consistent with implementation-science domains identified in prior reviews of clinical AI adoption [35,36].

**Figure 3.**
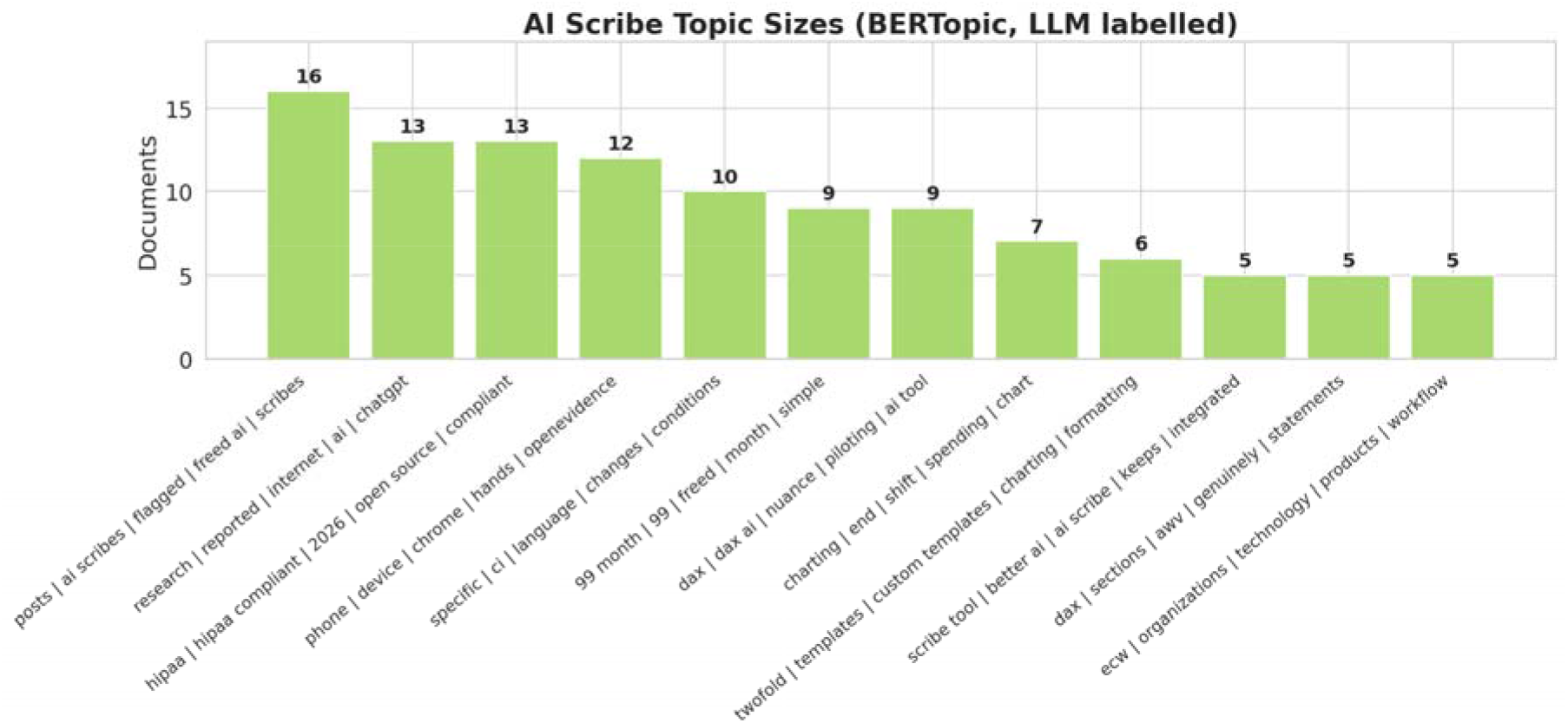
BERTopic theme sizes (number of posts per theme) after noise-pattern removal.

The intertopic distance map (Figure 4) confirmed semantic separation between vendor-comparison clusters (DAX, Heidi/templates, Freed/cost) and the more clinically framed clusters (research/ChatGPT, specific/conditions, charting/end-of-shift). HIPAA and compliance occupied a distinct region close to the research/internet cluster, consistent with the framing of compliance as an evidence-and-policy issue rather than a workflow issue [37].

**Figure 4.**
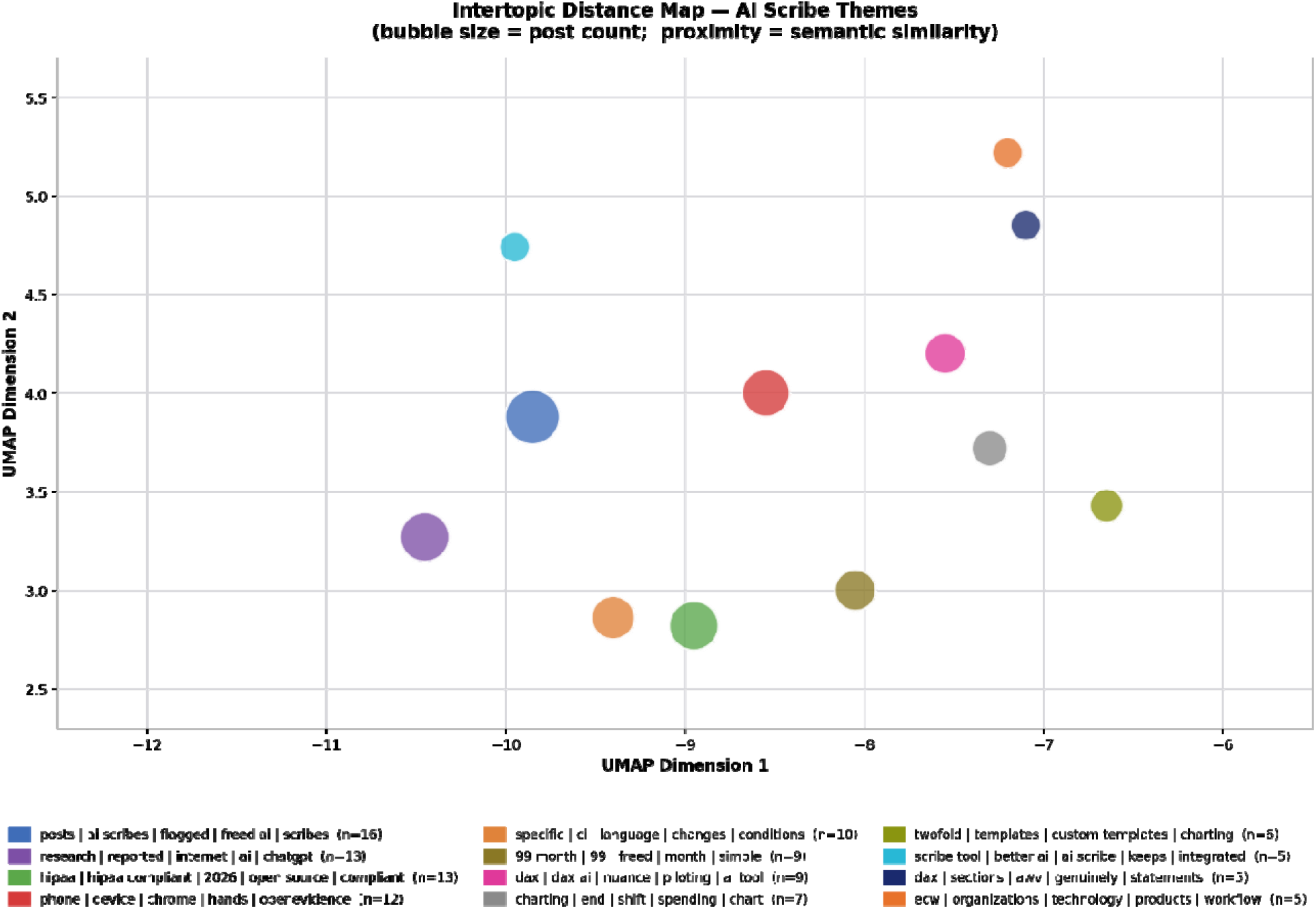
Intertopic distance map. Bubble size = post count; proximity = semantic similarity in UMAP-projected c-TF IDF space.

Per-theme c-TF-IDF keywords (Figure 5) provide the lexical fingerprint for each cluster [20]. Vendor-specific themes are anchored by named products (dax, dax ai, nuance, dragon, dictation; heidi, twofold, custom templates, formatting; freed, 99 month, simple, tier). The HIPAA cluster is structured around hipaa compliant, open source, ai medical, data, and policy. The phone/device cluster surfaces practical hardware constraints (phone, device, chrome, hands, mic, ambient scribe, app), which is operationally important for outpatient and inpatient use cases where dedicated workstations are not available [13].

**Figure 5.**
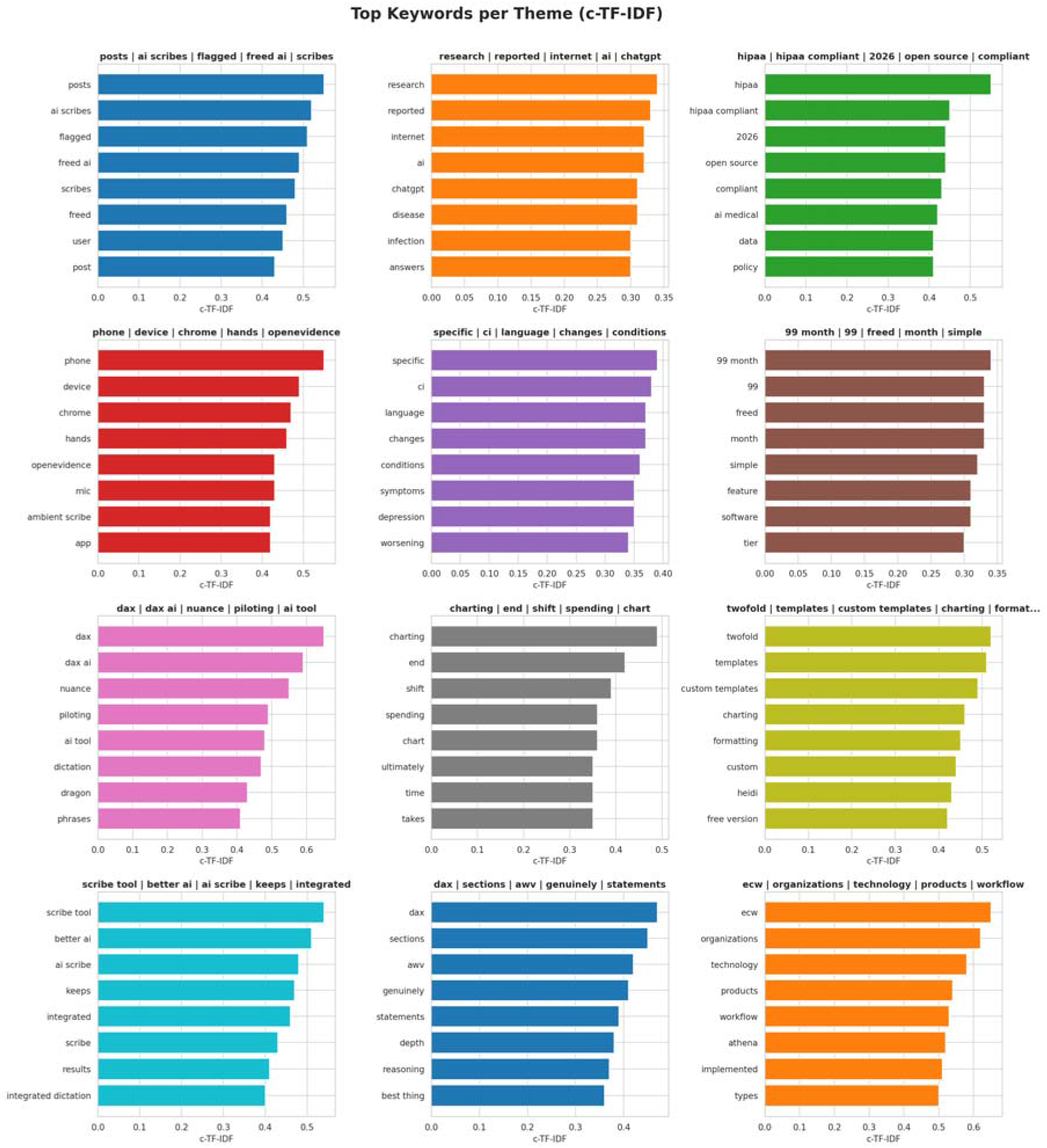
Top c-TF-IDF keywords for each of the twelve retained themes.

### 3.4 Sentiment distribution

Across the full corpus, sentiment was 61.4% neutral, 21.6% positive, and 17.0% negative (Figure 6). The dominance of neutral classifications is consistent with the problem-solving register of professional subreddits, where users predominantly seek and exchange operational information rather than express affect [16,24].

**Figure 6.**
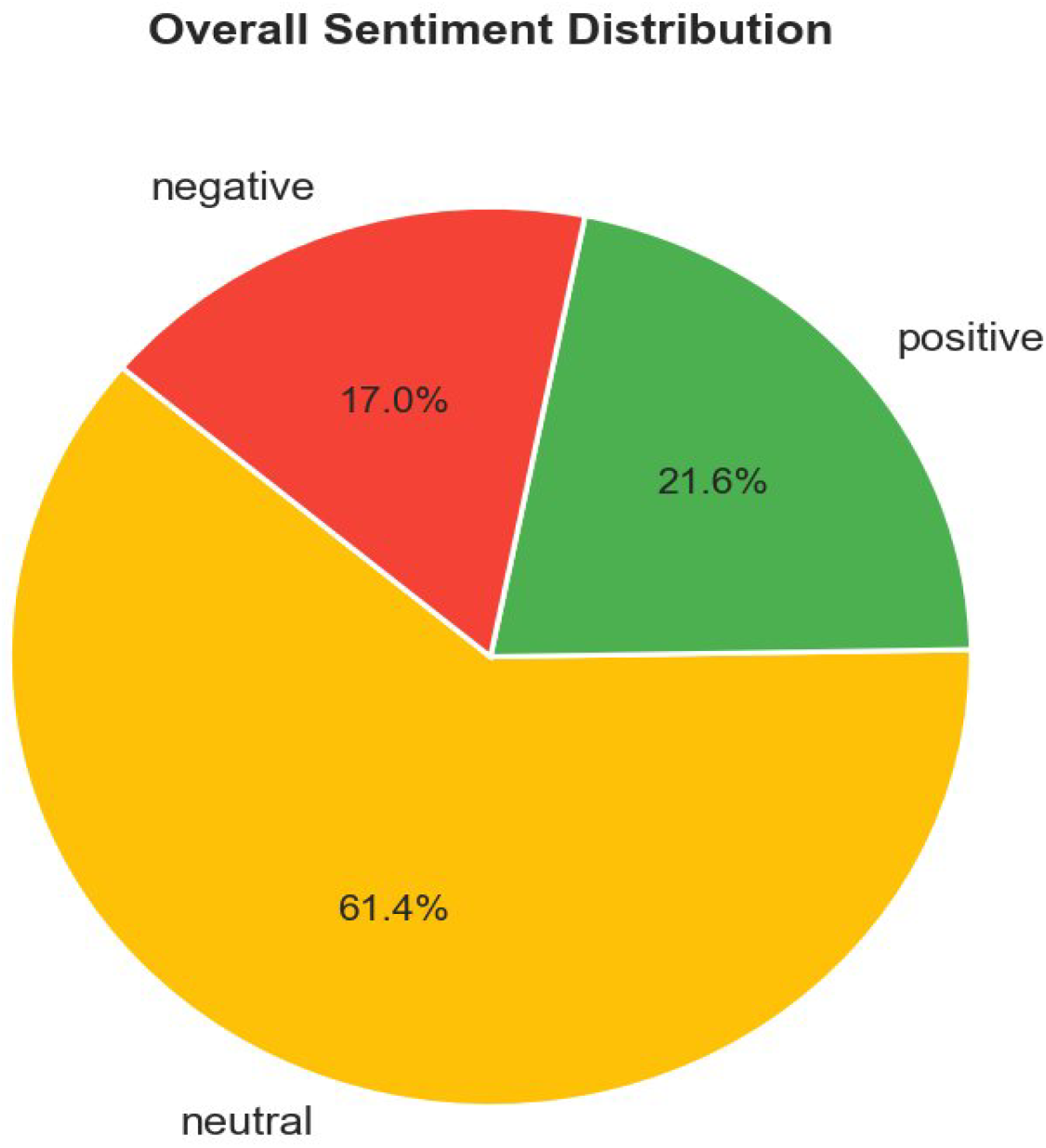
Overall sentiment distribution across 176 AI scribe posts.

Sentiment varied substantially by community (Figure 7). r/medicine showed the highest proportion of positive posts (about 40%) but also a high negative share (about 26%), consistent with polarised debate among practising physicians [38]. r/nursing was the most neutral (about 87%) and least positive (about 6%), suggesting a more wait-and-see stance, possibly reflecting limited exposure of nursing workflows to current ambient scribe products [39]. r/FamilyMedicine and r/physicianassistant showed broadly similar mixed profiles, while r/Psychiatry was strongly tilted toward neutrality with limited negativity.

**Figure 7.**
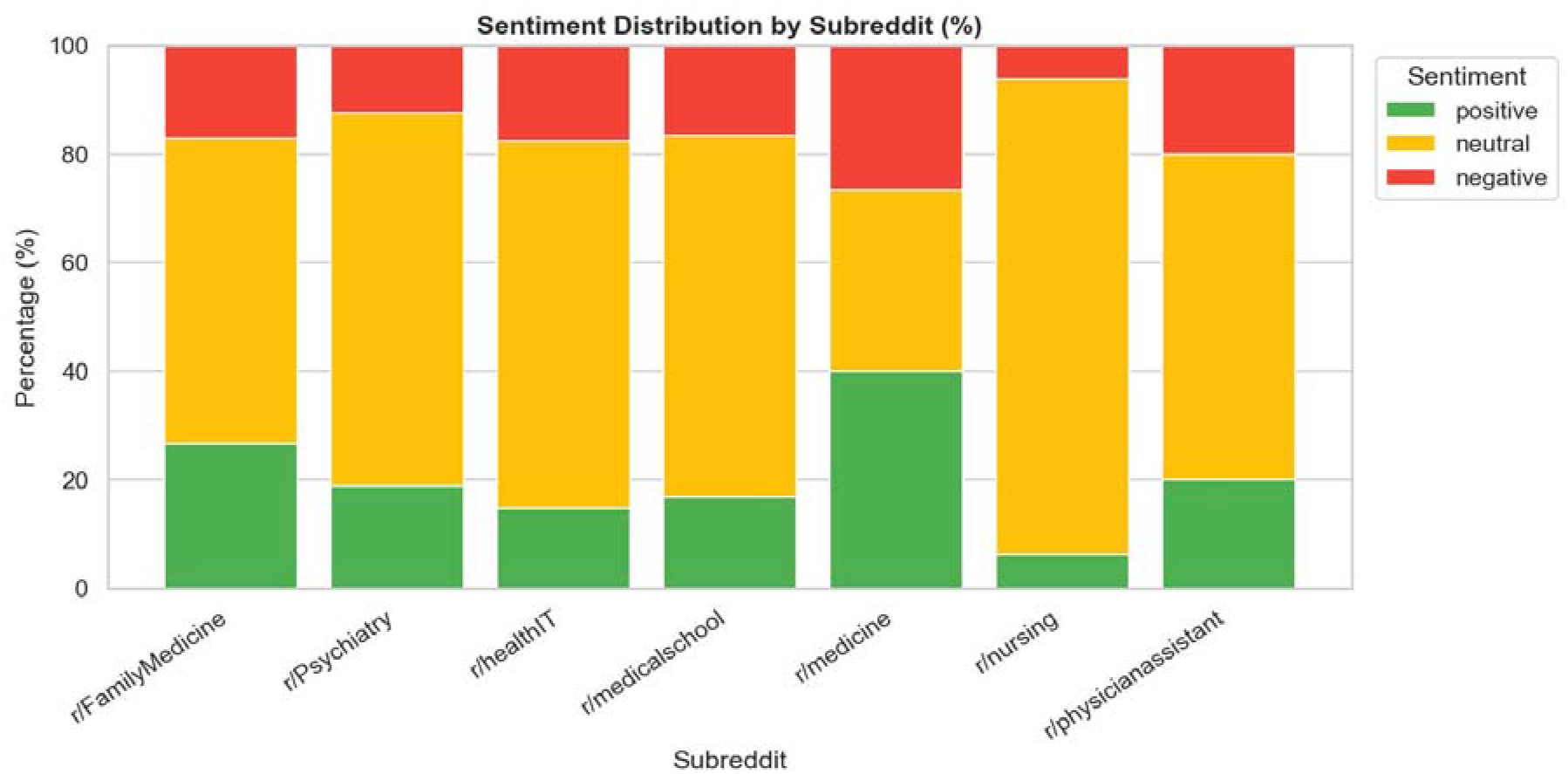
Sentiment distribution by subreddit (percentage). r/medicine shows the most polarised discourse; r/nursing is the most neutral.

### 3.5 Theme-level sentiment

Theme-level sentiment (Figure 8) revealed substantial heterogeneity. The DAX/Nuance/AI tools theme stood out as the most net-positive cluster, with approximately 55% positive and only 11% negative posts, reflecting strong endorsement among clinicians who have piloted DAX or Dragon Medical [7,8]. HIPAA and compliance discussion was also net-positive (about 31% positive, about 8% negative), suggesting that clinicians view existing compliance frameworks for major vendors as workable [37]. Conversely, the charting/end-of-shift theme was overwhelmingly neutral to negative (about 71% neutral, about 29% negative, 0% positive), capturing burnout-adjacent posts where AI scribes are discussed as a partial solution rather than a satisfying one [3,4]. The flagged Freed-scribes thread and the scribe-tool/better-AI/integrated cluster both carried elevated negative shares (about 37 to 40%), reflecting moderation friction and unmet expectations from early adopters [40].

**Figure 8.**
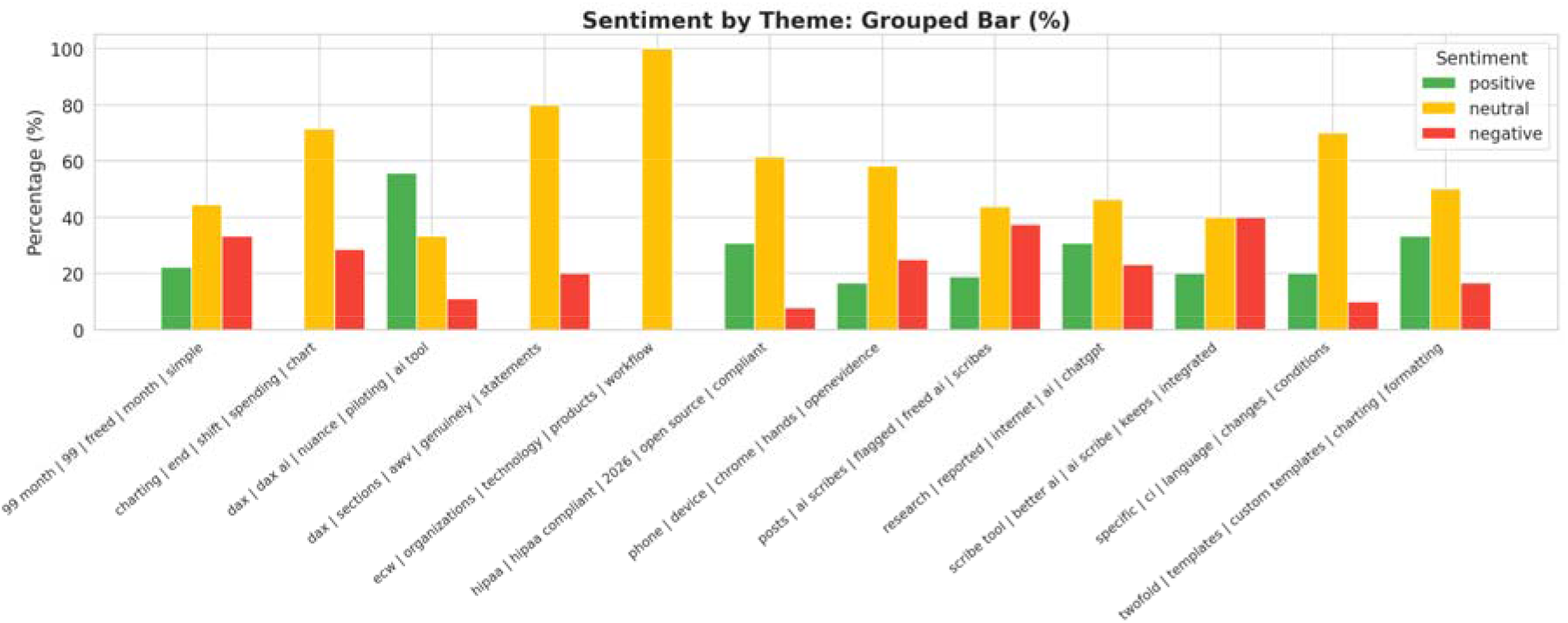
Sentiment composition by theme (grouped bars, percentage).

The two-dimensional theme-positioning plot (Figure 9) makes the favourable to unfavourable structure of the discourse explicit. Themes in the lower-right quadrant, namely high positive and low negative, are the unambiguous bright spots: DAX/Nuance/AI tools, HIPAA/compliance, and twofold/Heidi templates. Themes in the upper-left and upper-middle quadrants, namely scribe tool/better-AI/integrated, posts/AI-scribes/flagged, and charting/end-of-shift, represent areas where clinician concern outpaces enthusiasm. The eCW/organisations theme sits at the origin (zero positive, zero negative) because it consists entirely of neutral product-listing posts.

**Figure 9.**
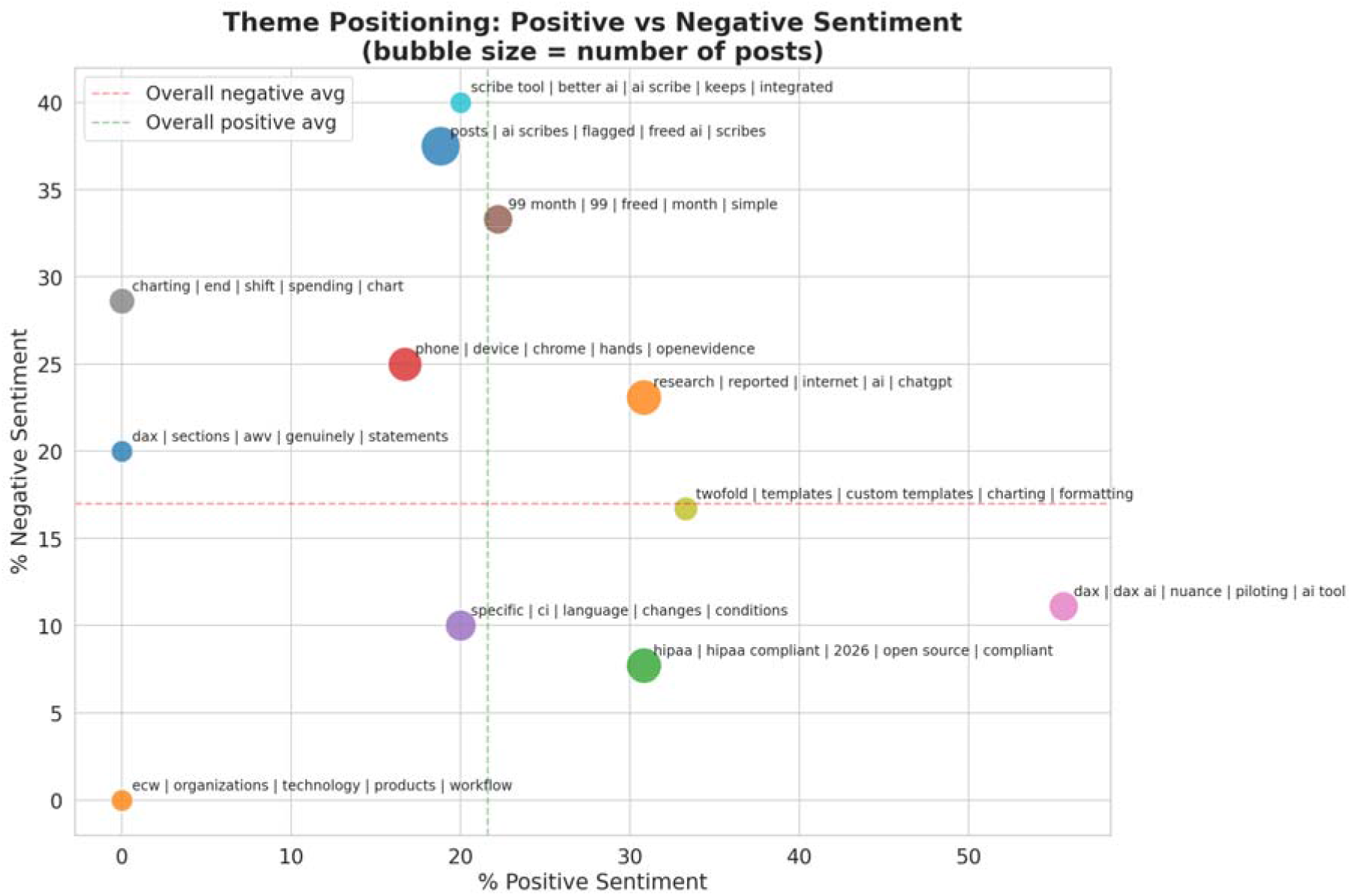
Theme positioning by positive vs negative sentiment. Bubble size = number of posts. Dashed lines mark the corpus-level averages.

Polarity distributions per theme (Figure 10) reinforce the same picture and add dispersion information: DAX/Nuance/AI tools shows a tight upper-tail distribution centred well above zero, while charting/end-of-shift is the only theme with a median polarity below zero. Most other themes are centred at neutrality with long tails toward both extremes, reflecting genuine disagreement among clinicians [38].

**Figure 10.**
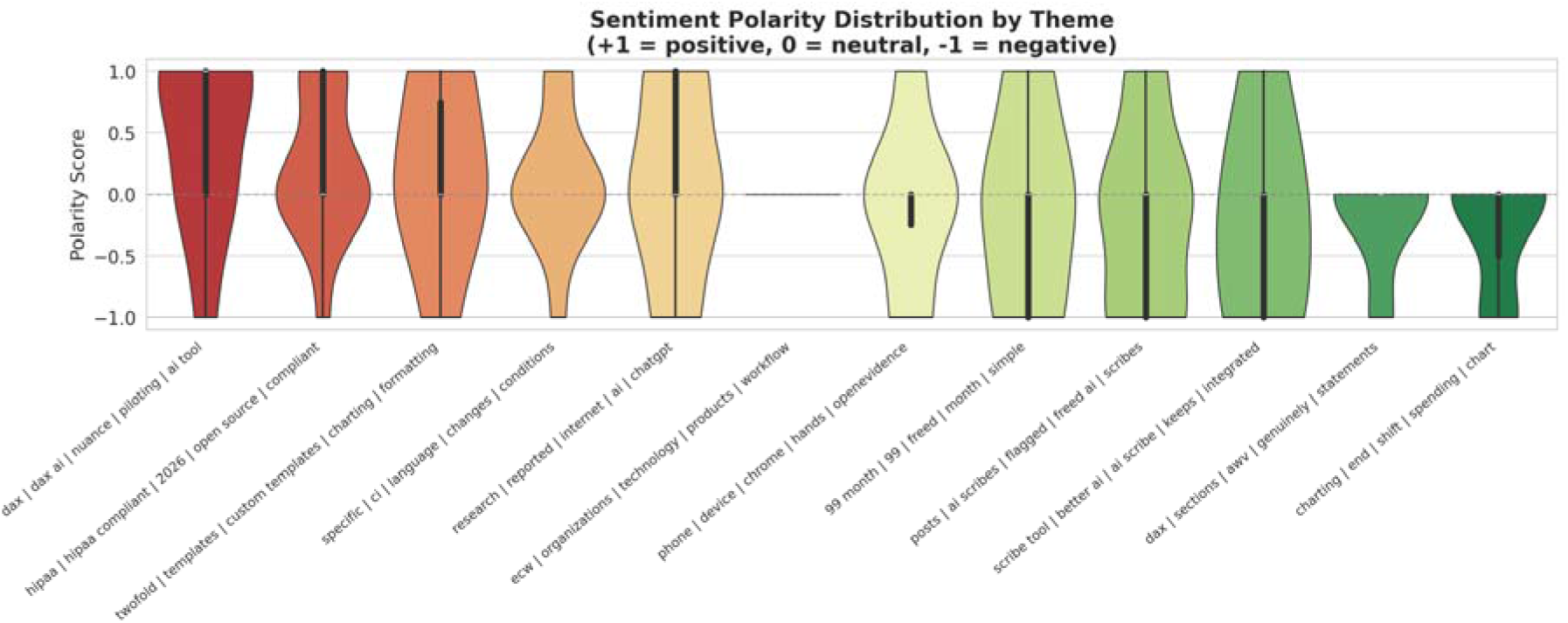
Sentiment polarity distribution by theme (+1 = positive, 0 = neutral, −1 = negative).

### 3.6 Engagement

Median upvote score and median comment count by theme (Figure 11) identify the topics that draw the most community attention [33]. The scribe-tool/better-AI/integrated and the research/ChatGPT themes attracted the highest median upvote scores (about 58 and about 44 respectively), while the Freed pricing and research themes drove the most comments (about 45 and about 41 medians). Pricing and product-comparison threads therefore not only carry sentiment signal but also dominate community engagement, reinforcing that practical adoption questions are the primary axis of clinician interest [11,12]. The eCW/organisations cluster, by contrast, was both small in volume and low in engagement, suggesting that organisational technology lists generate less substantive discussion than direct user experience.

**Figure 11.**
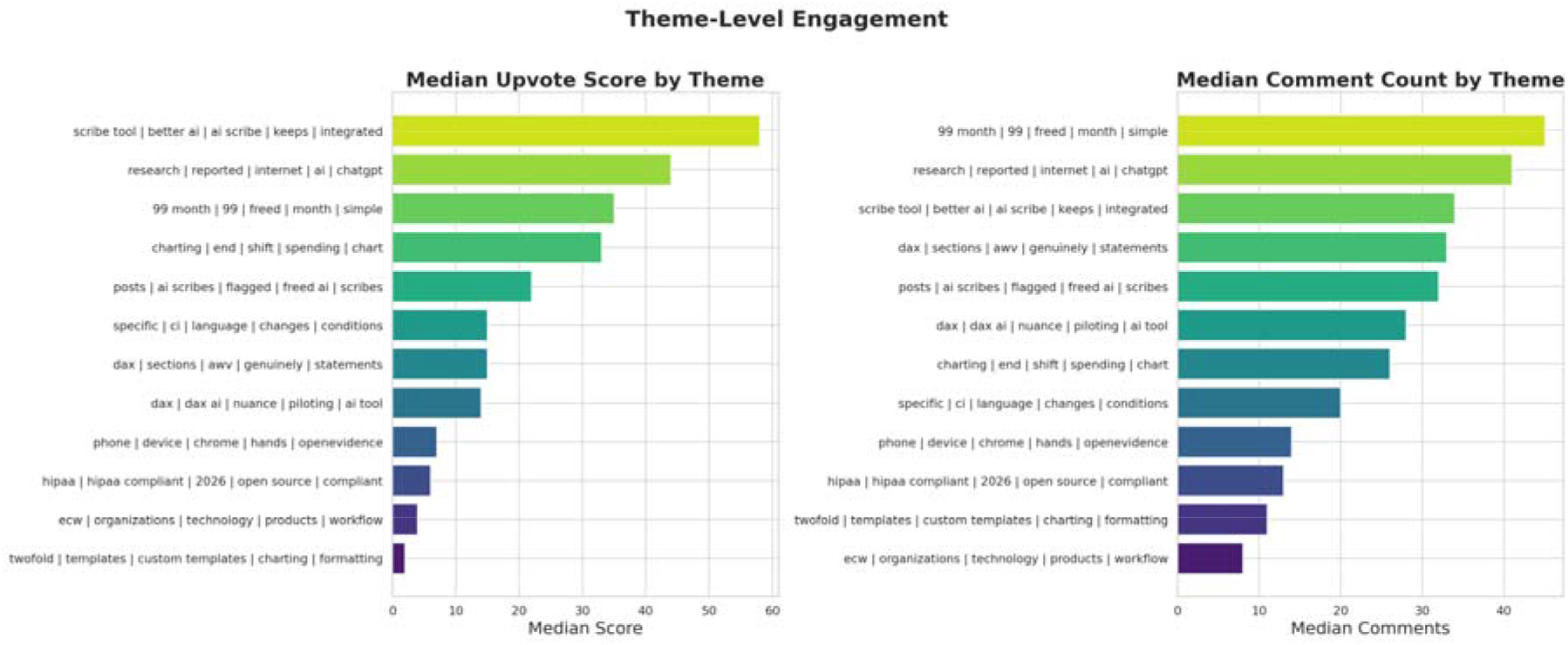
Theme-level engagement: median upvote score (left) and median comment count (right).

## 4. Discussion

### 4.1 Principal findings

This analysis provides one of the first systematic readings of clinician discourse on ambient AI scribes outside of vendor-sponsored or institution-led evaluations [10,11]. Three findings stand out. First, despite the high public profile of ambient documentation tools, the modal sentiment in clinician forums is neutral rather than enthusiastic, indicating that the dominant register is informational and operational rather than promotional [16,24]. Second, sentiment polarity is concentrated at the product level: DAX/Nuance and Heidi-style template tools are favourably discussed, whereas charting-fatigue and moderation-flagged threads carry negative valence [7,40]. Third, the issues that drive the most community engagement are pricing, vendor comparison, and EHR integration, not accuracy or hallucination, which appear far less prominently than the technology-policy discourse implies [12,41].

### 4.2 Implications for implementation

For health systems considering deployment of ambient AI scribes, the discourse maps cleanly onto established implementation-science domains [35,36]. Outer-setting concerns appear in the HIPAA and open-source clusters, where compliance and data-handling questions dominate [37]. Inner-setting and characteristics-of-the-individual concerns surface in the device, charting, and templates clusters, where workflow fit and learning curve are debated [42]. Innovation characteristics, including relative advantage, complexity, and cost, are most visible in the vendor-comparison and pricing clusters, where the Freed price point recurs as a benchmark [11]. Process-of-implementation concerns are less visible, suggesting that clinicians on Reddit are predominantly individual adopters rather than members of structured pilot programmes; this is itself a finding, in that grass-roots adoption may be running ahead of formal change-management [43].

### 4.3 Strengths and limitations

Strengths include the use of a transparent, reproducible pipeline combining state-of-the-art sentence embeddings, density-based clustering, and large-language-model thematic labelling [20,27,30], with explicit noise-pattern removal to guard against off-topic contamination. The two-tier keyword filter materially improved precision over single-tier filters used in earlier social-media studies of clinical AI [16,18].

Several limitations should be acknowledged. The corpus is small (n = 176) and reflects only English-language posts on Reddit, with selection bias toward US-based primary care and health-IT communities; nursing and inpatient perspectives are under-represented [39]. Sentiment was scored with a Twitter-trained model, which may misclassify long-form clinical reasoning; technical content with hedged language may be over-classified as neutral [22]. Topic modelling is sensitive to UMAP and HDBSCAN hyper-parameters, and the small corpus limits the granularity of themes that can be reliably recovered [21]. We did not analyse comments, where richer back-and-forth deliberation often occurs [33]. Finally, Reddit is a self-selected community and does not represent the broader population of clinicians; findings should be triangulated with survey and EHR-based usage data before being read as population-level estimates [14,15].

### 4.4 Future work

Three extensions are warranted. First, expanding to comment-level data and longer time windows would allow longitudinal tracking of vendor sentiment as new entrants and pricing models emerge. Second, cross-platform replication on physician-only forums, LinkedIn, and X would test the generalisability of the theme structure reported here [16,17]. Third, linking thematic clusters to objective adoption indicators, such as vendor-reported deployment counts, EHR app-marketplace listings, and published clinical evaluations, would convert this qualitative landscape into a quantitative diffusion model suitable for implementation-science evaluation [35,43].

## 5. Conclusion

Clinician discourse on ambient AI scribes across professional Reddit communities is best characterised as cautiously favourable, predominantly neutral, and operationally focused. Twelve coherent themes were recovered, with vendor comparison, cost, HIPAA compliance, and EHR integration emerging as the most engaged-with issues. Sentiment is concentrated at the product level rather than the category level, with DAX/Nuance and template-based tools eliciting the most positive responses, and burnout-adjacent charting threads the most negative. These findings can directly inform implementation strategy, vendor benchmarking, and policy guidance for ambient documentation tools, while aligning with qualitative evidence on clinician and nurse experiences highlighting workflow integration, usability, and contextual adoption challenges [44,45], and demonstrate the value of transparent NLP pipelines applied to clinician-generated online discourse as a complement to formal evaluation.

## Data Availability

All data produced in the present work are contained in the manuscript

## Declarations

### Data availability

All Reddit posts analysed were publicly available at the time of retrieval. The full analysis pipeline, including scraping, filtering, topic modelling, and visualisation code, is available from the corresponding author on reasonable request. No personally identifiable information is reported.

### Ethics

The study used only public, non-personally-identifiable posts and did not require institutional ethics review.

### Author contributions

RS conceived the study, designed and implemented the pipeline, performed the analysis, generated all figures, and wrote the manuscript. QX validated.

